# Dynamic Contrast Enhanced MRI Mapping of Vascular Permeability for Evaluation of Breast Cancer Neoadjuvant Chemotherapy Response Using Image-to-Image Conditional Generative Adversarial Networks

**DOI:** 10.1101/2024.09.04.24313070

**Authors:** Chad A. Arledge, Alan H. Zhao, Umit Topaloglu, Dawen Zhao

## Abstract

Dynamic contrast enhanced (DCE) MRI is a non-invasive imaging technique that has become a quantitative standard for assessing tumor microvascular permeability. Through the application of a pharmacokinetic (PK) model to a series of T1-weighed MR images acquired after an injection of a contrast agent, several vascular permeability parameters can be quantitatively estimated. These parameters, including K_trans_, a measure of capillary permeability, have been widely implemented for assessing tumor vascular function as well as tumor therapeutic response. However, conventional PK modeling for translation of DCE MRI to PK vascular permeability parameter maps is complex and time-consuming for dynamic scans with thousands of pixels per image. In recent years, image-to-image conditional generative adversarial network (cGAN) is emerging as a robust approach in computer vision for complex cross-domain translation tasks. Through a sophisticated adversarial training process between two neural networks, image-to-image cGANs learn to effectively translate images from one domain to another, producing images that are indistinguishable from those in the target domain. In the present study, we have developed a novel image-to-image cGAN approach for mapping DCE MRI data to PK vascular permeability parameter maps. The DCE-to-PK cGAN not only generates high-quality parameter maps that closely resemble the ground truth, but also significantly reduces computation time over 1000-fold. The utility of the cGAN approach to map vascular permeability is validated using open-source breast cancer patient DCE MRI data provided by The Cancer Imaging Archive (TCIA). This data collection includes images and pathological analyses of breast cancer patients acquired before and after the first cycle of neoadjuvant chemotherapy (NACT). Importantly, in good agreement with previous studies leveraging this dataset, the percentage change of vascular permeability K_trans_ derived from the DCE-to-PK cGAN enables early prediction of responders to NACT.

## 1. Introduction

Neoadjuvant chemotherapy (NACT) has been established as a standard of care for patients with locally advanced breast cancer and has been expanded to include patients with early-stage breast cancer^1^. The primary goal of NACT is to downstage the tumor prior to surgery, enabling improved operability and conservation of healthy breast tissue^2-5^. Pathologic complete response (pCR) to NACT has been shown to be prognostic for survival^1^. However, only 6-25% of breast cancer patients experience pCR to NACT^3^. Moreover, pathologic analysis must be performed after multiple cycles of NACT and surgery, limiting the ability to assess patient responses early in the treatment process. Currently, there is significant interest in non-invasive means to predict the pathologic response status to NACT prior to surgery, enabling clinicians to adjust personalized treatment regimens based on early responses, which in turn could lead to improved patient outcomes^2^.

To this end, various quantitative imaging modalities have been investigated for assessment of pathologic response status in breast cancer patients receiving NACT attributable to their non-invasive ability to assess tumor biology^3,5,6^. Early changes in underlying tumor biology characteristics including permeability, perfusion, metabolism, oxygenation, and cell density could enable early prediction of pathological response. In recent years, researchers have identified dynamic contrast enhanced (DCE) MRI and its resulting pharmacokinetic (PK) vascular permeability parameters as a potential biomarker for early assessment of NACT pathological response in breast cancer patients^2,3,7^. Tudorica et al. have shown that percentage changes in DCE MRI vascular permeability parameters are good to excellent predictors of pCR to NACT, with univariate logistic regression (ULR) c-statistic values ranging from 0.804-0.967 in a cohort of 28 invasive ductal carcinoma patients^2^. Namely, early changes in K_trans_, a measure of capillary permeability, immediately following the first cycle of NACT has shown the greatest promise for early prediction of pathologic response.

Despite its potential, the use of DCE MRI for monitoring treatment response has several limitations hindering its widespread clinical implementation. First, there is significant variability in image acquisition protocols and scanners across different institutions, which can lead to non-reproducible and inconsistent results in clinical oncology. Standardization of DCE MRI acquisition will enhance the reproducibility and consistency of DCE MRI studies. Moreover, DCE MRI requires sophisticated software for solving complex PK mathematical models that are used to generate vascular permeability parameters. There is also significant variation in not only PK models, but also the software tools/packages used to solve these models. It is well recognized that commercialization of a standardized software tool for generating vascular permeability parameters in DCE MRI will help establish the widespread clinical implementation of DCE MRI^7^. Furthermore, establishment of a simple software tool for generating vascular permeability parameters will enhance the ease of implementation and decrease the complexity and time required to solve these mathematical models.

To this end, several research groups including ours have investigated deep learning as a potential tool to standardize PK analysis and reduce computational complexity. These studies have applied deep learning approaches for quantification of vascular permeability in applications including brain cancer^8-10^, stroke^11^, pancreatic cancer^12^, and head and neck cancer^13^. The deep learning approaches explored in these studies include 1D convolutional neural network (CNN), 2D CNN, and recurrent neural networks (RNNs) such as gated recurrent unit (GRU) and long short-term memory (LSTM) networks. Although these studies have demonstrated the utility of deep learning for efficient quantification of vascular permeability, it is still uncertain if deep learning can reliably monitor tumor vascular permeability treatment responses with the same level of precision as conventional methods. Moreover, with the recent surge in the development of novel deep learning approaches, several innovative techniques have emerged, offering potential improvements in both accuracy and robustness for quantifying vascular permeability in DCE MRI.

One of these novel deep learning approaches, generative adversarial network (GAN), specifically image-to-image conditional GAN (cGAN), has proven to be a robust approach in computer vision for complex cross-domain translation tasks^14,15^. During training of an image-to-image cGAN, a Generator neural network learns to create synthetic images that are similar to a target domain using images from another domain. Simultaneously, a Discriminator neural network evaluates the synthetic images against real images and determines if they are real or fake, guiding the Generator to produce increasingly realistic images over time. By leveraging this sophisticated adversarial training scheme between two neural networks, image-to-image cGANs can generate high-quality images that closely resemble the ground truth. Several studies have revealed the robust capabilities of cGANs for complex medical imaging tasks, including synthetic image generation^16,17^, image de-noising^18^, segmentation^19,20^, super-resolution^21^, and modality transfer^22^. Specifically, image-to-image cGANs have recently been successfully applied to challenging cross-domain translation tasks in medical imaging, such as synthesizing apparent diffusion coefficient (ADC) maps from diffusion-weighted imaging^23^ and mapping cerebral blood volume (CBV) from standard MRI scans^24^.

Clearly, image-to-image cGANs are an attractive approach for translation of DCE MRI to PK vascular permeability parameter maps. However, to the best of our knowledge, an image-to-image cGAN has not been previously investigated for DCE-to-PK cross-domain translation tasks. In the present study, we have developed a novel DCE-to-PK cGAN approach for quantification of vascular permeability parameter maps in DCE MRI. The utility of the deep learning approach is validated in open-source breast cancer patient DCE MRI data provided by The Cancer Imaging Archive (TCIA). This data collection contains patient images and pathological analyses acquired before and after the first cycle of NACT from a multi-center data analysis challenge^7,25^. Moreover, the NACT induced early changes in vascular permeability as quantified by the DCE-to-PK cGAN has been correlated with pathological treatment response.

## 2. Materials and Methods

### 2.1 Patient Data

An open-source collection of breast cancer DCE MRI data provided by the Quantitative Imaging Network (QIN) and published on TCIA was used in the current study for training, testing, and validating the proposed image-to-image cGAN approach^7,25^. The data collection was used in a previous study investigating the response of breast cancer patients to NACT in a multi-QIN center data analysis challenge. As described previously^7^, 20 DCE MRI scans were acquired across ten breast cancer patients. Each patient was subject to two imaging exams, one acquired prior to the start of treatment, V1, and another acquired after the first cycle of treatment, V2. MRI studies were performed on a Siemens 3T TIM Trio system. DCE MRI was acquired using a 3D gradient echo-based time-resolved angiography with stochastic trajectories (TWIST) sequence (TR/TE: 6.2/2.9 msec; flip-angle: 10 degrees; field of view: 30 to 34 cm; in-plane matrix size: 320 × 320; slice thickness: 1.4 mm; parallel imaging acceleration factor: 2; temporal resolution: 18 to 20 sec). Following the acquisition of two baseline images, a contrast agent Gd(HP-D03A) [ProHance] was injected intravenously (0.1 mmol/kg at 2 ml/s) with a programmable power injector, followed by a 20 ml saline flush.

In addition to DCE MRI data, average tumor baseline longitudinal relaxation rate R_1_ (R_10_), population-averaged arterial input function (AIF), tumor region of interest (ROI), and pathologic response status were provided for each patient for PK modeling and analysis. As described previously, tumor average R_10_ values were determined by comparing the signal intensity of baseline DCE images without contrast to spatially registered proton density images acquired immediately before DCE MRI^26^. The provided population-averaged AIF was obtained in a previous sagittal breast DCE MRI study, which employed the same contrast agent injection protocol^27^. Patient tumor ROIs were drawn by experienced breast radiologists at Oregon Health and Science University on post-contrast DCE MRI. Pathologic status was determined through pathologic comparisons pre-NACT and following the final cycle of NACT. A binary classification of pCR or non-pCR (pathologic nonresponse and pathologic partial response) was provided for each patient^7^.

### 2.2 DCE MRI Preprocessing

DCE MRI scans were cropped to isolate patient tumor-bearing breasts and remove background anatomical structures including the lungs and heart, resulting in an in-plane matrix size of 160 × 160. The scans were truncated to twenty-eight frames to allow for all imaging datasets having the same temporal size. DCE images were then motion corrected using an in-house MATLAB co-registration algorithm.

### 2.3 Pharmacokinetic Modeling

Dynamic signal intensity changes in DCE MRI and patient-specific R_10_ values were used to determine C_t_, the dynamic concentration of contrast within the tissue, according to the following equations:

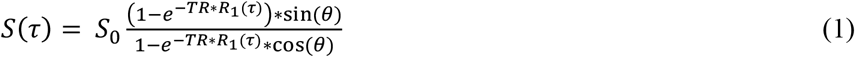

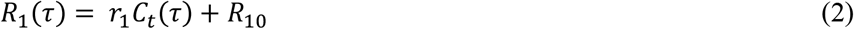

where S and S_0_ is the DCE MRI signal intensity with and without contrast agent, respectively, θ is the flip angle of the DCE MRI sequence, and r_1_ is the relaxivity of the contrast agent. Following C_t_ mapping, the Extended Tofts Model (ETM) was applied to estimate vascular permeability parameters according to Eqn. 3:

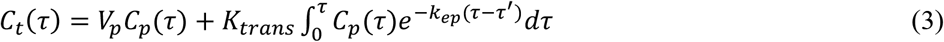

where C_p_ is the dynamic concentration of contrast within the plasma, also known as the AIF. V_p_, K_trans_, and k_ep_ are vascular permeability parameters that describe the fractional volume of blood plasma, the transfer rate of contrast agent from the blood plasma to the extravascular extracellular space (EES), and the reverse transfer rate of the contrast agent from the EES to the blood plasma, respectively^10,28,29^. A non-linear least squares (NLLS) curve-fitting algorithm was used to map the PK parameters to C_t_ and C_p_ for each voxel. The bounds for curve-fitting of each vascular parameter were: K_trans_ = [1e^-8^, 2], k_ep_ = [1e^-8^, 2], and V_p_ = [1e^-8^, 0.2]. All mathematical modeling algorithms were implemented in MATLAB R2024a with an Intel Xeon CPU @ 3.70 GHz and 32 GB RAM.

### 2.4 DCE-to-PK cGAN Architecture

The proposed cGAN architecture for translating DCE MRI to PK vascular permeability parameters is shown in **Figure 1**. Patient-specific R_10_ values were broadcasted to a matrix size of 160 × 160 and concatenated with corresponding DCE MRI patient scans. The Generator neural network translates R_10_ + DCE MRI into synthetic vascular permeability K_trans_ maps. The Discriminator neural network determines if the input K_trans_ maps from the ETM or the Generator are real or fake, conditioned on corresponding R_10_ + DCE MRI. In the current study, we used a dual-pathway CNN from our previous work^9,10^ for the Generator and a conditional PatchGAN for the Discriminator^14^.

**Figure 1.**
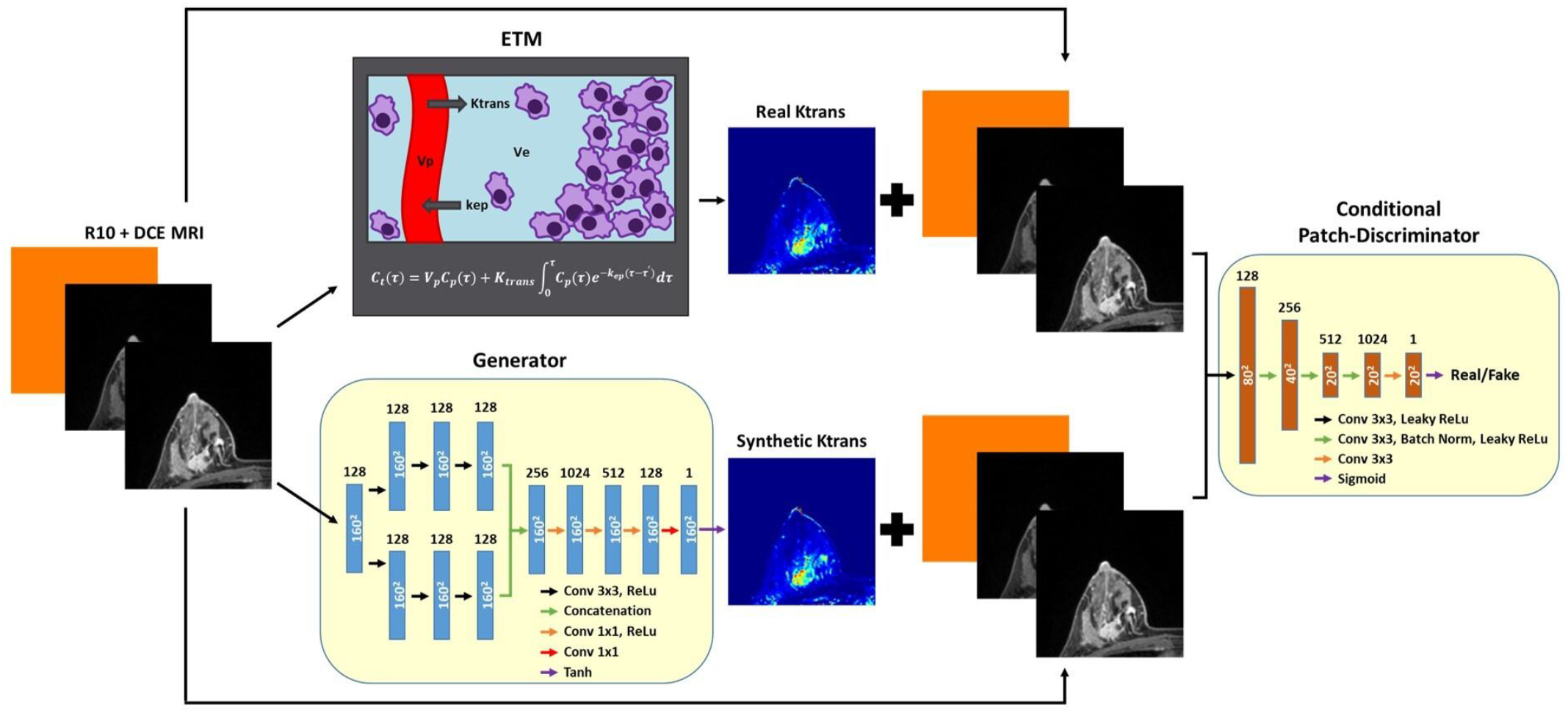
Pathway for DCE-to-PK cGAN training. A Generator neural network was trained to translate input R_10_ + DCE MRI to synthetic K_trans_ maps. Simultaneously, a Discriminator neural network was trained to distinguish between the Generator synthetic K_trans_ maps and real ETM K_trans_ maps, conditioned on input R_10_ + DCE MRI. A custom least squares patch-based adversarial loss incorporating an L1-loss term was used to update both network parameters until the resulting synthetic K_trans_ maps generated by the Generator appeared indistinguishable from real ETM K_trans_ maps.

For the Generator, an image input layer and convolutional layer were followed by the two parallel pathways, local and global, to capture multi-scale features. The local pathway consists of three convolutional non-dilated layers. The global pathway consists of three convolutional layers that were dilated by factors of 2, 4, and 8, respectively. The dual-pathway convolutional layers were designed with 128 filters with a size of 3 × 3. Local and global pathways were then concatenated and followed by four fully connected convolutional layers of 1024, 512, 128, and one filter, each with a size of 1 × 1. A ReLU activation layer followed each convolutional layer, except for the final convolutional layer, which was instead followed by a Tanh layer. Zero padding was applied to every convolutional operation.

For the Discriminator, a conditional PatchGAN was implemented to down-sample K_trans_ maps by a factor of eight and discriminate based on patches. The Discriminator was conditioned with corresponding R_10_ + DCE MRI used to create the real and synthetic K_trans_ maps. An image input layer was followed by a fully convolutional architecture with five convolutional layers of 128, 256, 512, 1024, and one filter, each with a size of 3 × 3. Batch normalization was applied to all convolutional layers, except for the first and final convolutional layer. A leaky ReLU activation layer followed each convolutional layer, apart from the final convolutional layer, which was instead followed by a Sigmoid layer.

### 2.5 Model Training

DCE MRI and corresponding K_trans_ maps were normalized to [0 1] for model training. K-fold cross validation was performed to allow all patients to serve as testing data of the DCE-to-PK cGAN. To this end, imaging data from a single patient (n = 240 DCE slices) from both pre and post-NACT (V1 and V2) were isolated and saved for network testing. The remaining patient data (n = 2,160 DCE slices) were then randomly split into an 80:20 training to validation ratio. Using the Adam optimizer, hyper-parameters were manually adjusted to achieve the best network performance with minimum error: a learning rate of 1e^-5^ for both the Generator and Discriminator, mini-batch size of 16, and maximum epochs of 200. Training data was shuffled at every epoch. A custom least squares cGAN-based approach^30^ incorporating an L1-loss (mean absolute error) component was implemented for the Generator and Discriminator loss functions as follows:

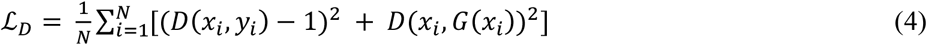

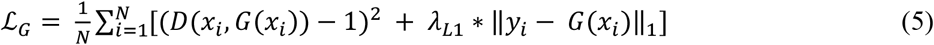

where N is the mini-batch size, x_i_ is the conditioned R_10_ + DCE MRI, y_i_ is the ground-truth ETM K_trans_ maps, *G*(*x*_*i*_) is the Generator synthetic K_trans_ maps, and λ_L1_ is the weight of the L1 loss term, which was set to 1. Following training, the patient testing dataset was fed directly into the Generator to produce synthetic K_trans_ maps that were then rescaled back to their original distribution. This process was repeated 10-fold for all patients. All neural network algorithms were implemented in MATLAB R2024a with an NVIDIA GeForce RTX 3090.

### 2.6 Statistical Analysis

Patient intratumoral individual pixel, slice mean, and tumor mean K_trans_ for both V1 and V2 were quantified using tumor ROI provided by the TCIA. Linear regression was applied to individual patients as well as an ensemble of patients K_trans_ values for statistical correlation and significance. The root-mean-squared error (RMSE), mean absolute error (MAE), normalized RMSE (nRMSE), normalized MAE (nMAE), Pearson R-squared (R^2^), and concordance correlation coefficient (CCC) were determined for each patient pixel-by-pixel K_trans_ between the DCE-to-PK cGAN and ETM. nRMSE and nMAE were normalized using patient standard deviation (SD) of target ETM K_trans_. ULR models were fit to compute the c-statistic, the area under the Receiver Operating Characteristic (ROC) curve, for V1 K_trans_, V2 K_trans_, and percentage K_trans_ change from V1 to V2 for both the ETM and cGAN approaches. Two-tailed and unpaired Student’s t-tests were used to determine statistical significance between pre- and post-NACT. *p* < .05 was considered to indicate statistical significance. Data were presented as mean ± SD.

## 3. Results

The DCE-to-PK cGAN was found to generate vascular permeability parameter maps in breast cancer TCIA patient datasets (n = 120 slices) in 2.5 seconds on average with an NVIDIA GeForce RTX 3090, significantly lowering the average computation time over 1000-fold relative to the ETM (150 min average on Intel Xeon CPU @ 3.70 GHz and 32 GB RAM). A representative non-pCR patient to NACT is depicted in **Figure 2**. DCE MRI revealed a large, highly enhanced tumor mass as well as multiple infiltrative lesions. These tumors were enhanced at both pre-treatment, V1, and post-NACT, V2. Vascular permeability K_trans_ maps produced by the cGAN closely resembled ETM counterparts, exhibiting excellent spatial correlation and high structural similarity (**Figure 2a**). This patient was found to have elevated vascular permeability K_trans_ at V1 (ETM = 0.0233 ± 0.0035 min^-1^; cGAN = 0.0230 ± 0.0035 min^-1^) that was largely retained at V2 (ETM = 0.0213 ± 0.0058 min^-1^; cGAN = 0.0200 ± 0.0053 min^-1^), with excellent agreement between the ETM and cGAN (**Figure 2b**). Linear regression (**Figure 2c and 2d**) revealed a significant linear correlation (p < 0.0001) between the cGAN and ETM for both slice-based mean K_trans_ (n = 43; R^2^ = 0.98) and pixel-by-pixel K_trans_ (n = 20,873; R^2^ = 0.98) for this patient.

**Figure 2.**
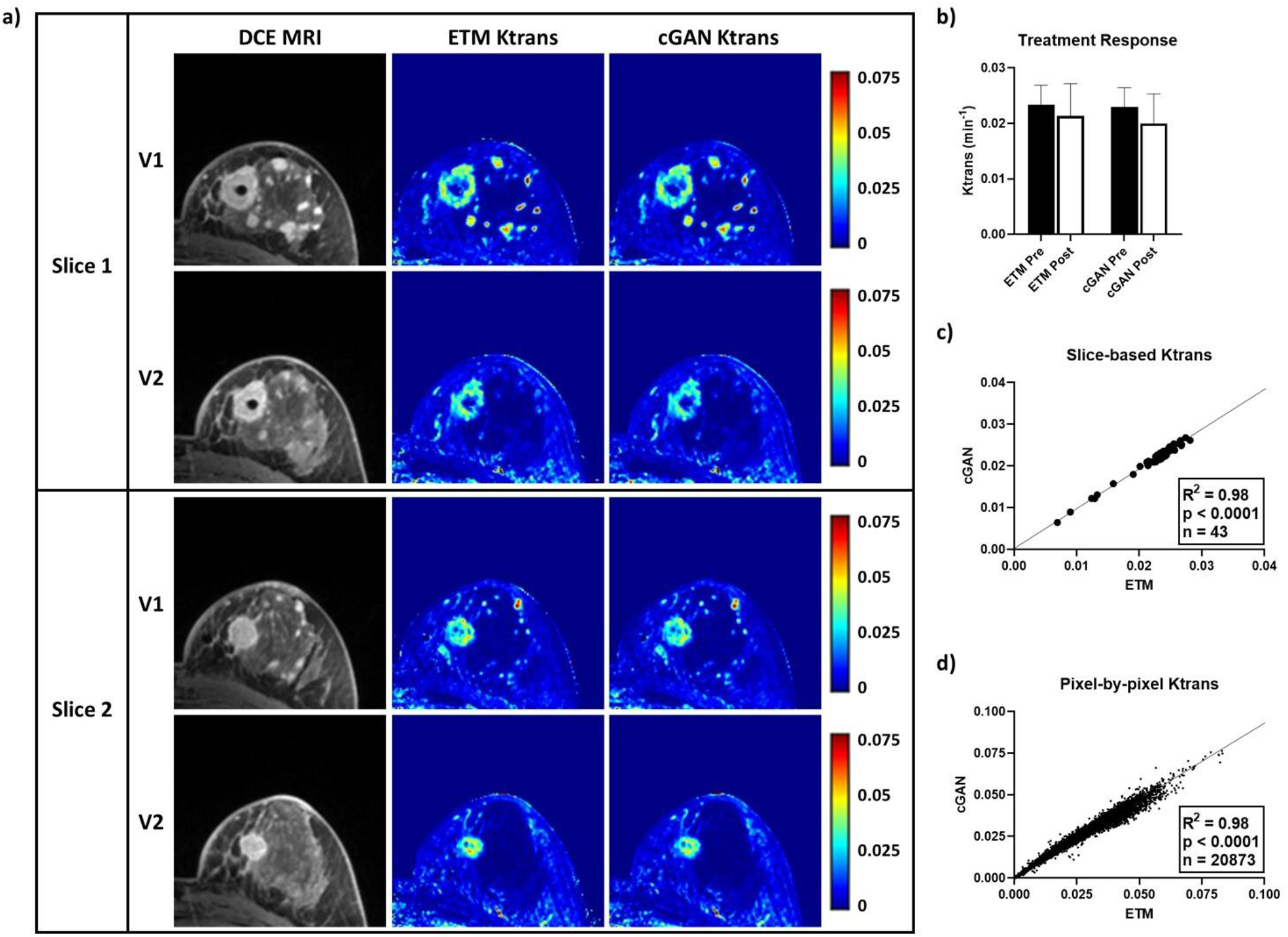
TCIA non-pCR patient tested using the DCE-to-PK cGAN. Two representative DCE MRI slices from a non-pCR patient revealed highly enhanced tumor lesions at both pre-treatment (V1) and post-NACT (V2). Notably, ETM and cGAN K_trans_ maps showed excellent spatial correlation and high structural similarity (**a**). Quantification of tumor mean K_trans_ at V1 (n = 22 slices) and V2 (n = 21 slices) revealed a small decrease in vascular permeability at V2 by both the ETM and cGAN (**b**). Analysis of slice-based mean K_trans_ (**c**) and pixel-by-pixel K_trans_ (**d**) showed strong correlations (R^2^ = 0.98) between the cGAN and ETM (p < 0.0001). Mean ± SD.

Similar to the non-pCR patient, as shown in **Figure 3**, DCE MRI of a representative pCR patient revealed a large, highly enhanced tumor at both V1 and V2. ETM and cGAN K_trans_ maps were almost indistinguishable, exhibiting excellent spatial correlation and high structural similarity (**Figure 3a**). While the patient was found to have elevated vascular permeability K_trans_ at V1 (ETM = 0.0383 ± 0.0164 min^-1^; cGAN = 0.0380 ± 0.0160 min^-1^), the patient had markedly decreased K_trans_ at V2 (ETM = 0.0155 ± 0.0016 min^-1^; cGAN = 0.0156 ± 0.0016 min^-1^) in good agreement by the ETM and cGAN (**Figure 3b**). Slice-based mean K_trans_ (n = 31; R^2^ = 0.99) and pixel-by-pixel K_trans_ (n = 3,031; R^2^ = 0.99) revealed a significant linear correlation (p < 0.0001) between the cGAN and ETM for this patient (**Figure 3c and 3d**). Clearly, the DCE-to-PK cGAN accurately monitored vascular permeability responses in non-pCR (**Figure 2**) and pCR (**Figure 3**) patients with high structural similarity to the ETM.

**Figure 3.**
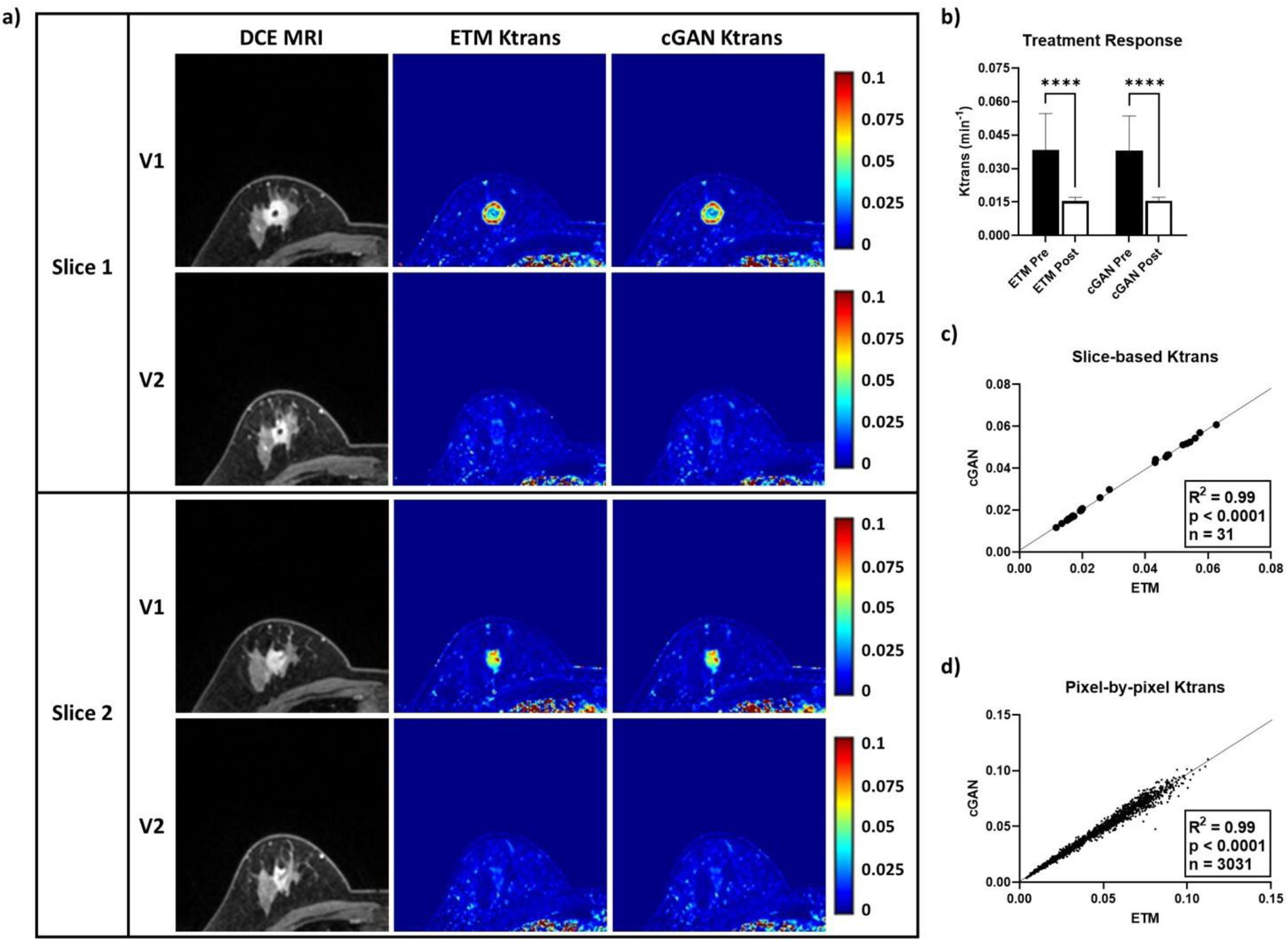
TCIA pCR patient tested using the DCE-to-PK cGAN. Two representative DCE MRI slices from a pCR patient revealed a highly enhanced tumor lesion at both pre-treatment, V1, and post-NACT, V2. Notably, K_trans_ maps produced by the ETM and cGAN had excellent spatial correlation and high structural similarity (**a**). Quantification of tumor mean K_trans_ (**b**) at V1 (n = 19 slices) and V2 (n = 12 slices) revealed substantially decreased vascular permeability at V2 by both the ETM and cGAN (p < 0.0001). Analysis of slice-based mean K_trans_ (**c**) and pixel-by-pixel K_trans_ (**d**) showed strong correlations (R^2^ = 0.99) between the cGAN and ETM (p < 0.0001). Mean ± SD, ****p < 0.0001.

The DCE-to-PK cGAN deciphered intra-patient heterogeneous tumor responses in good agreement to the ETM, further demonstrating its utility (**Figure 4**). Anatomical DCE images of a representative breast cancer patient revealed two tumor lesions, lesion 1 (green arrow) and lesion 2 (red arrow). Both lesions were largely enhanced at V1 and V2 (**Figure 4a**). ETM and cGAN maps showed elevated vascular permeability K_trans_ at V1 for lesion 1 (ETM = 0.0247 ± 0.0090 min^-1^; cGAN = 0.02451 ± 0.0091 min^-1^) and lesion 2 (ETM = 0.0283 ± 0.0077 min^-1^; cGAN = 0.0287 ± 0.0078 min^-1^). In response to NACT, lesion 1 had markedly decreased K_trans_ (**Figure 4b**; ETM = 0.0104 ± 0.0030 min^-1^; cGAN = 0.0102 ± 0.0032 min^-1^), whereas lesion 2 retained elevated permeability (**Figure 4c**; ETM = 0.0232 ± 0.0051 min^-1^; cGAN = 0.0239 ± 0.0050 min^-1^). Importantly, these heterogeneous responses between the two lesions were recapitulated with high accuracy by the cGAN. Moreover, there was significant linear correlation (p < 0.0001) between the cGAN and ETM for both slice-based mean K_trans_ (n = 28; R^2^ = 0.99) and pixel-by-pixel K_trans_ (n = 6,471; R^2^ = 0.98) for this patient (**Figure 4d and 4e**).

**Table 1.** Predictive performance of DCE-to-PK cGAN.

| Patient | RMSE | MAE | nRMSE | nMAE | R<sup>2</sup> | CCC |
| --- | --- | --- | --- | --- | --- | --- |
| Patient 01 | 1.98E-3 | 1.28E-3 | 0.167 | 0.108 | 0.981 | 0.985 |
| Patient 05 | 2.63E-3 | 1.62E-3 | 0.113 | 0.069 | 0.988 | 0.993 |
| Patient 06 | 1.15E-3 | 7.70E-4 | 0.181 | 0.120 | 0.973 | 0.985 |
| Patient 08 | 1.04E-3 | 5.37E-4 | 0.100 | 0.052 | 0.990 | 0.995 |
| Patient 10 | 2.18E-3 | 1.33E-3 | 0.264 | 0.161 | 0.947 | 0.968 |
| Patient 12 | 3.19E-3 | 1.11E-3 | 0.316 | 0.110 | 0.954 | 0.959 |
| Patient 13 | 1.95E-3 | 1.38E-3 | 0.223 | 0.157 | 0.981 | 0.978 |
| Patient 14 | 1.97E-3 | 1.26E-3 | 0.138 | 0.088 | 0.982 | 0.991 |
| Patient 15 | 1.06E-3 | 6.01E-4 | 0.114 | 0.065 | 0.987 | 0.994 |
| Patient 16 | 2.46E-3 | 1.63E-3 | 0.176 | 0.116 | 0.973 | 0.985 |

**Figure 4.**
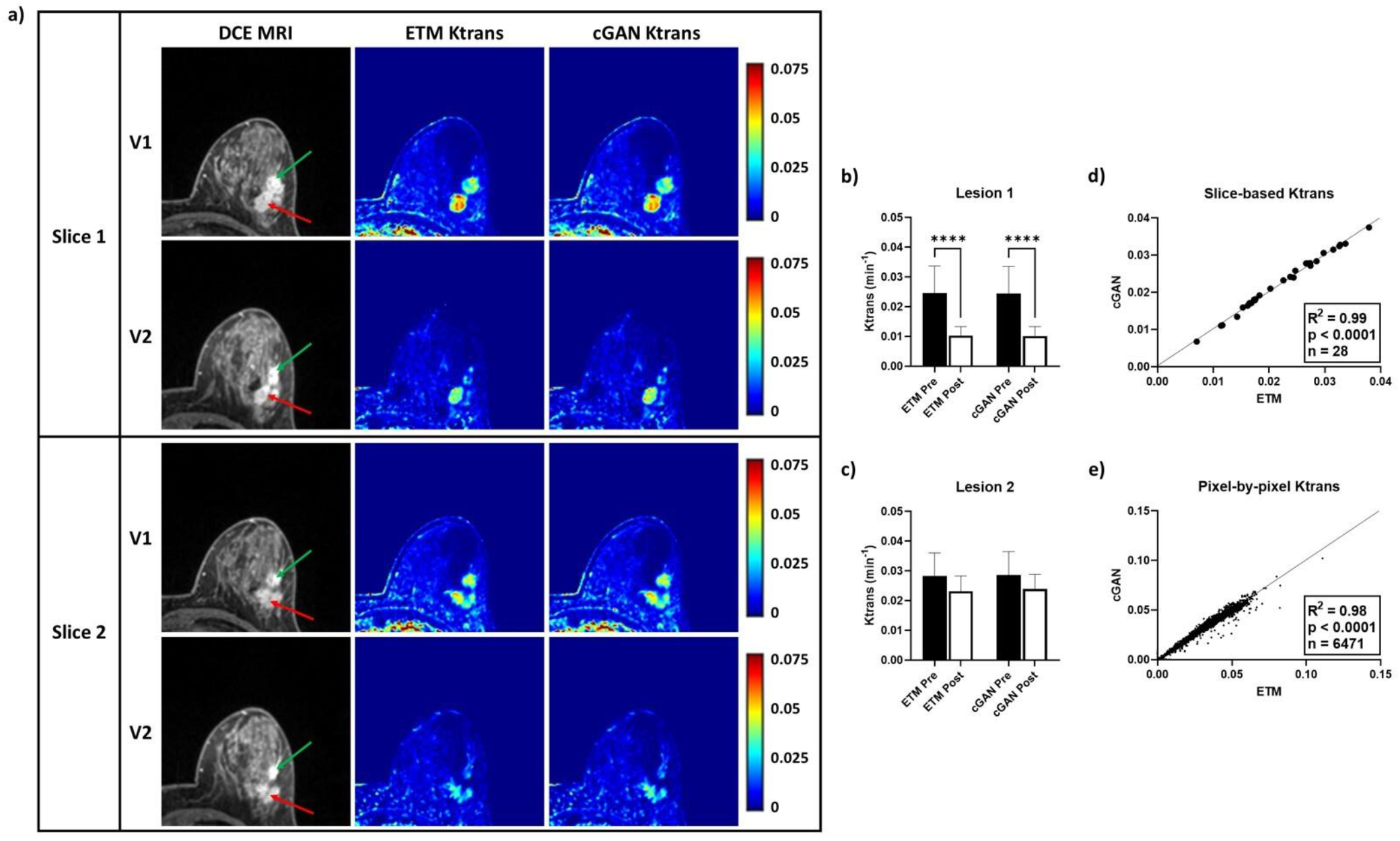
DCE-to-PK cGAN deciphered intra-patient heterogeneous tumor response. In a representative patient, two highly enhanced tumor lesions, lesion 1 (green arrow) and lesion 2 (red arrow), were identified on DCE images (**a**). Notably, ETM and cGAN K_trans_ maps showed elevated vascular permeability at V1 for both lesions. Interestingly, both ETM and cGAN V2 K_trans_ maps revealed markedly decreased vascular permeability for lesion 1 (**b**) and retained vascular permeability for lesion 2 (**c**). Slice-based mean K_trans_ (**d**) and pixel-by-pixel K_trans_ (**e**) analysis showed strong correlations (R^2^ = 0.98) between the cGAN and ETM (p < 0.0001). Mean ± SD, ****p < 0.0001.

Error analysis (**Table 1**) across all patient’s intratumoral pixel-by-pixel K_trans_ values showed low RMSE and MAE for the cGAN. Importantly, all patients had less predictive error than the SD seen on ETM maps (nRMSE and nMAE < 1). R^2^ and CCC revealed strong correlations between the cGAN and ETM approaches for quantifying K_trans_ (R^2^ and CCC > 0.9). As shown in **Figure 5**, an intratumoral ensemble of tumor mean K_trans_ (n = 20), slice-based mean K_trans_ (n = 359), and pixel-by-pixel K_trans_ (n = 143,640) across all patients demonstrated significant linear correlations (p < 0.0001) between the ETM and cGAN (R^2^ ≥ 0.98). Negligent over or under-prediction by the cGAN for tumor mean, slice-based mean, and pixel-by-pixel K_trans_ was found from Bland-Altman plot analysis (not shown here).

**Figure 5.**
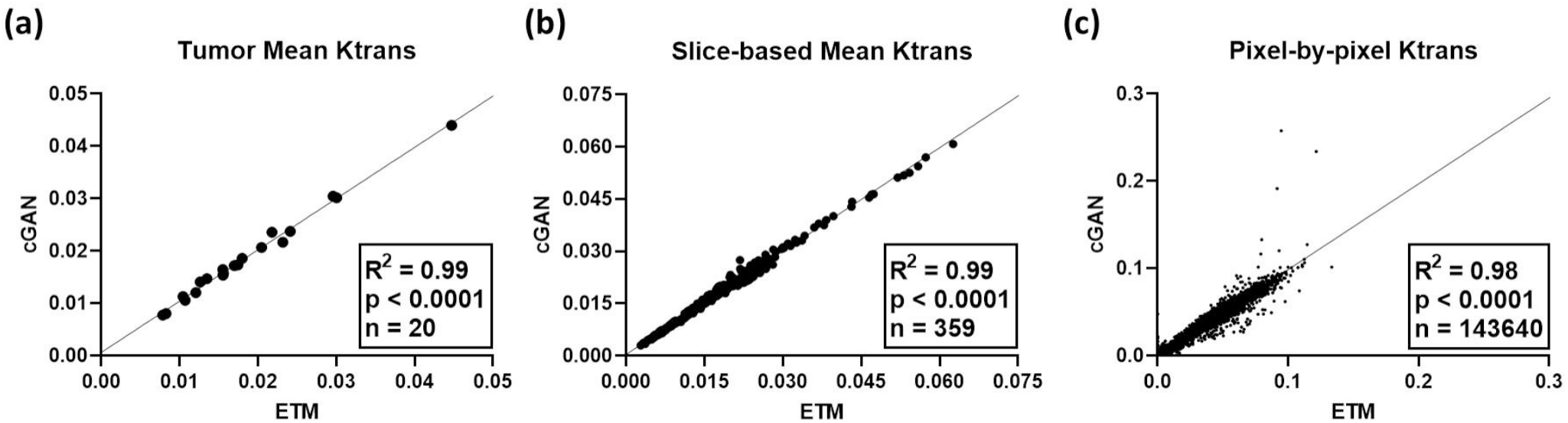
Patient ensemble linear regression analysis. Tumor mean K_trans_ (**a**) across all patients at both V1 and V2 (n = 20) revealed a significant linear correlation between the ETM and cGAN (p < 0.0001; R^2^ = 0.99). Similarly, intratumoral slice-based mean K_trans_ (**b**) and pixel-by-pixel K_trans_ (**c**) had strong linear correlations between the ETM and cGAN with R^2^ = 0.99 (n = 359) and 0.98 (n = 143,640), respectively (p < 0.0001).

Pathological evaluations of individual patient responses to NACT were performed previously by Oregon Health & Science University in a study by Huang et al^7^. In this group of patients, seven were identified as non-pCR and three were pCR. Based on these classifications, we plotted tumor mean K_trans_ at V1 and V2 as well as percentage K_trans_ change between V1 and V2 (**Figure 6**). Clearly, scatter plots of tumor mean K_trans_ for both the ETM (**Figure 6a**) and cGAN (**Figure 6b**) showed differential PK vascular permeability responses to NACT between non-pCR (red triangles) and pCR patients (black circles). The pCR patients had a more prominent decrease in tumor mean K_trans_ after the first cycle of NACT in comparison to non-pCR patients. There was great agreement in K_trans_ values and percentage K_trans_ change between the cGAN and ETM. Importantly, the DCE-to-PK cGAN quantified percentage K_trans_ change clearly distinguished non-pCR from pCR patients in good agreement to the ETM (**Figure 6c**). Furthermore, concurring with previous work leveraging this dataset^7^, ULR models found percentage K_trans_ change was an excellent predictor of pathological response (**Table 2**).

**Table 2.** ULR c-statistic values for early prediction of pathologic response to NACT.

| Model | V1 $K_{trans}$ | V2 $K_{trans}$ | %$K_{trans}$ Change |
| --- | --- | --- | --- |
| ETM | 0.619 | 0.857 | 1.00 |
| cGAN | 0.619 | 0.857 | 1.00 |

**Figure 6.**
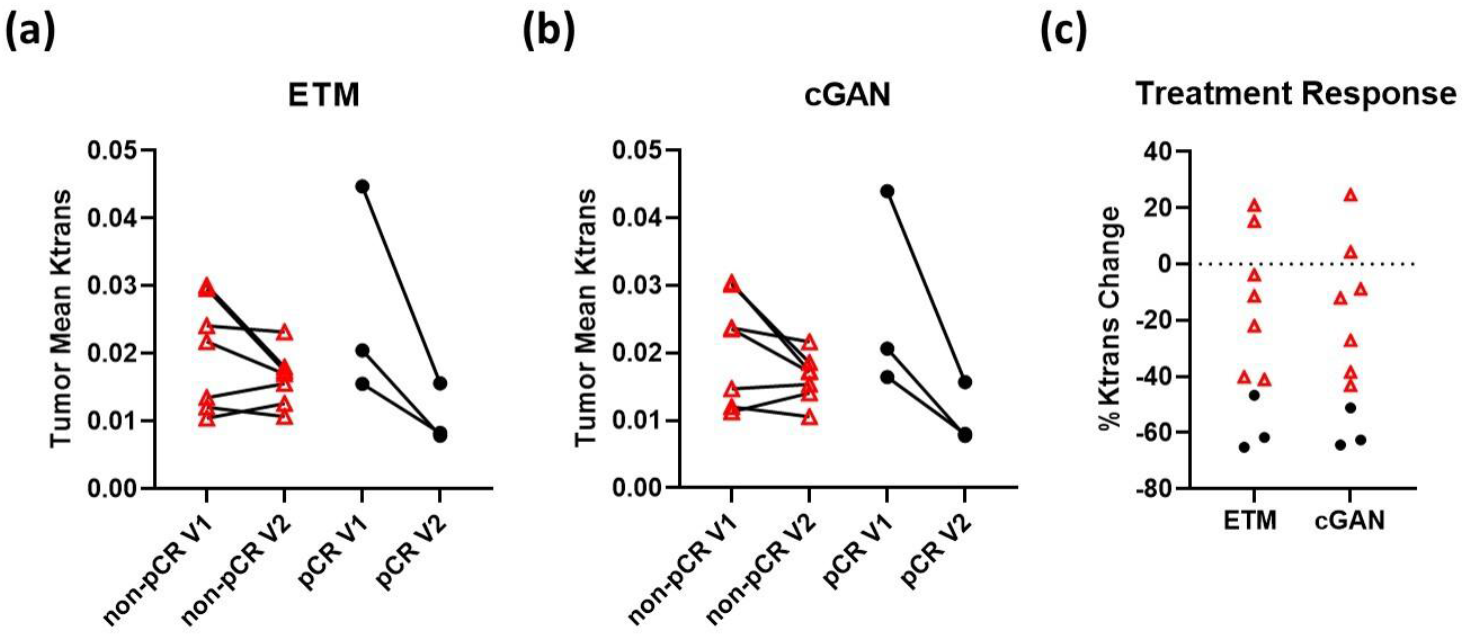
DCE-to-PK cGAN enables early prediction of responders to NACT. Scatter plots of tumor mean K_trans_ for both ETM (**a**) and cGAN (**b**) revealed differential vascular changes to NACT between non-pCR patients (red triangles) and pCR patients (black circles). Notably, in good agreement with previous work^7^, K_trans_ percentage changes (**c**) clearly distinguished the non-pCR from the pCR patients.

## 4. Discussion

DCE MRI is becoming increasingly common in standard-of-care cancer imaging regimens attributable to its noninvasive ability to assess tumor biology in the form of tumor microvascular permeability. However, DCE MRI has several limitations that still hinder its widespread clinical implementation. These limitations include variability in image acquisition protocols, the need for standardized PK analysis techniques, and the requirement for advanced computational tools. Addressing these issues could significantly enhance the clinical utility of DCE MRI in oncology. To this end, we have developed a novel DCE-to-PK cGAN approach for translation of DCE MRI to PK vascular permeability parameter maps.

An important finding from the present study is that the DCE-to-PK cGAN significantly lowered the computational time over 1000-fold relative the ETM, while generating synthetic K_trans_ maps that closely resembled real ETM K_trans_ maps (**Figures 2-4**). In addition to the excellent spatial correlation and visual structural similarity between cGAN and ETM maps, we found low intratumoral error (nMAE and nRMSE < 1.0) and strong correlation (R^2^ and CCC > 0.9) across all patients (**Table 1**). Furthermore, an ensemble of tumor-bearing K_trans_ values for all patients (**Figure 5**) revealed a significant linear correlation (p < 0.0001) for tumor mean, slice-based mean, and pixel-by-pixel K_trans_ (R^2^ ≥ 0.98). Clearly, the DCE-to-PK cGAN can efficiently quantify vascular permeability in good agreement to conventional PK models.

Several previous studies have identified DCE MRI derived vascular permeability parameters as a potential biomarker for early assessment of NACT pathological response in breast cancer patients^2,3,7^. These studies have suggested that NACT-induced tumor function changes, particularly vascular function changes, precede changes in tumor size, which is the current standard of care for evaluating treatment response^2,3,7^. Furthermore, these early changes in vascular function are prognostic for pathological response and may serve as a useful surrogate for early monitoring of treatment response. Indeed, as shown in **Figure 2 and 3**, representative non-pCR and pCR patients from TCIA data collection as used in this study have minimal change in tumor size following the first cycle of NACT. In good agreement with previous work of this data collection^7^, the non-pCR patient was found to retain elevated vascular permeability based on the ETM, whereas the pCR patient exhibited a significant decrease in vascular permeability parameter K_trans_ (p < 0.0001). Importantly, the DCE-to-PK cGAN correctly quantified these early vascular permeability changes in both non-pCR and pCR patients in excellent agreement to the ETM.

It is noteworthy that tumors in both non-pCR (**Figure 2**) and pCR (**Figure 3**) breast cancer patients were highly enhanced on DCE MRI at both V1 and V2, indicating lesions were permeable both pre-and post-NACT. However, as discussed above, non-pCR and pCR patients showed differential vascular permeability K_trans_ changes in response to NACT. K_trans_ is related to the rate of enhancement on DCE MRI scans, indicating that the rate of enhancement may be more important for monitoring treatment response than the presence of high contrast enhancement. Another noteworthy finding is that the DCE-to-PK cGAN not only accurately quantified vascular permeability changes between individual patients, but also accurately deciphered intra-patient heterogeneous tumor responses, as evidenced in the patient shown in **Figure 4**. We anticipate the DCE-to-PK cGAN can serve as a useful tool for monitoring both inter-patient and intra-patient tumor vascular permeability responses, which can aid in clinical decision-making and personalized medicine.

In a previous work leveraging this data collection by Huang et al., a multi-QIN center data analysis challenge revealed significant variability of vascular permeability parameters derived from twelve different DCE MRI computational modeling algorithms^7^. The authors found up to a two-fold to four-fold parameter value difference between software tools, albeit controlling for tumor ROI definition as well as R_10_ and AIF measurements. A separate study similarly revealed a 10-fold to 100-fold difference in vascular permeability parameters derived from different software tools in DCE MRI^31^. These differences in vascular permeability parameters based on different PK models and software tools could lead to non-reproducible results in DCE MRI studies across different institutions. Huang’s study showed strong agreement in percentage change of vascular permeability parameter K_trans_ for nearly all software tools, of which 11 out of 12 were good (0.8 ≤ c-statistic ≤ 0.9) to excellent (c-statistic ≥ 0.9) predictors of pathologic response by ULR analysis^7^.

In the present study leveraging the same dataset from Huang’s study^7^, we found our in-house ETM software returned smaller vascular permeability K_trans_ values than the majority of the other computational modeling algorithms used in the data analysis challenge. Our vascular permeability K_trans_ values produced by in-house ETM software were similar to the BWH-3D Slicer TM software tool^7,32^ used in the data analysis challenge (**Figure 6a**). The DCE-to-PK cGAN approach also predicted smaller vascular permeability K_trans_ values relative to the other software tools as the deep learning approach was trained to recapitulate our ETM software (**Figure 6b**). Nevertheless, percentage K_trans_ changes for both our ETM software and cGAN are in excellent agreement to all software tools as used in Huang’s study (**Figure 6c**)^7^. Similarly, we show that both our ETM software and cGAN approach are excellent predictors of pathologic response (c-statistic = 1.0) when considering percentage change of vascular permeability parameter K_trans_ (**Table 2**).

As revealed in the present study, an image-to-image cGAN may serve as a useful tool in DCE MRI for monitoring treatment response. However, it is worth noting that the current sample size of the study cohort is small. Including additional training and testing cases will further elucidate the long-term utility of the DCE-to-PK cGAN approach. To this end, with the recent surge in the development of novel artificial intelligence (AI) approaches, specifically generative AI, it would be interesting to evaluate the ability of these approaches to generate synthetic DCE MRI. Such synthetic data could be used to augment and expand training cases, potentially improving the performance of the DCE-to-PK cGAN. Moreover, a direct comparison of the DCE-to-PK cGAN to previously developed deep learning algorithms for efficient DCE MRI mapping was not performed in the current study but is warranted. It would similarly be interesting to evaluate an ensemble learning approach to combine the knowledge of alternative deep learning frameworks, which may lead to enhanced predictive accuracy over implementation of a single deep learning model.

## 5. Conclusion

In summary, we have developed a novel image-to-image cGAN approach for efficient translation of DCE MRI to PK vascular permeability parameter maps. The DCE-to-PK cGAN can generate vascular permeability parameter maps that closely resemble real parameter maps and reduce the computational time over 1000-fold compared to conventional computational tools. Importantly, percentage change of vascular permeability parameter K_trans_ maps derived from the cGAN approach were excellent early predictors of pathologic response in breast cancer patients to NACT, highlighting its utility for monitoring treatment response. We anticipate the DCE-to-PK cGAN will serve as a useful tool to standardize PK analysis and reduce computational complexity in both general oncologic imaging and clinical trials.

## Acknowledgements

The authors thank the Quantitative Imaging Network and The Cancer Imaging Archive for providing the open-source breast cancer DCE MRI data. We are grateful to Janet Zhang, Department of Radiology at The University of North Carolina School of Medicine, for valuable support and guidance interpreting DCE MRI scans.

## Data Availability

The dataset used in the present study are publicly available through The Cancer Imaging Archive^7,25^.

